# Is the BioNTech-Pfizer COVID-19 vaccination effective in elderly populations? Results from population data from Bavaria, Germany

**DOI:** 10.1101/2021.08.19.21262266

**Authors:** Delphina Gomes, Andreas Beyerlein, Katharina Katz, Gabriele Hoelscher, Uta Nennstiel, Bernhard Liebl, Klaus Überla, Rüdiger von Kries

## Abstract

**Background:** The effect of the BioNTech-Pfizer BNT162b2 vaccination in the elderly (≥80 years) could not be fully assessed in the BioNTech-Pfizer trial due to low numbers in this age group. We aimed to evaluate the effectiveness of the BioNTech-Pfizer (BNT162b2) vaccine to prevent SARS-CoV-2 infection and severe outcomes in octo- and novo-generians in a German state setting.

**Methods and Findings:** A prospective observational study of 708,187 persons aged ≥80 years living in Bavaria, Germany, was conducted between Jan 9 to Apr 11, 2021. We assessed the vaccine efficacy (VE) for two doses of the BNT162b2 vaccine with respect to SARS-CoV-2 infection and related hospitalisations and mortality. Additionally, differences in VE by age groups ≥80 to ≤89 years and ≥90 years were studied. Analyses were adjusted by sex.

By the end of follow-up, 63.8% of the Bavarian population ≥80 years had received one dose, and 52.7% two doses, of the BNT162b2 vaccine. Two doses of the BNT162b2 vaccine lowered the proportion of SARS-CoV-2 infections and related outcomes, resulting in VE estimates of 68.3% (95% confidence interval (CI) 65.5%, 70.9%) for infection, 73.2% (95% CI 65.3%, 79.3%) for hospitalisation, and 80.1% (95% CI 80.0%, 89.0%) for mortality. Sex differences in the risk of COVID-19 outcomes observed among unvaccinated persons disappeared after two BNT162b2 vaccine doses. Overall, the BNT162b2 vaccine was equally efficacious in octo- and novo-genarians.

**Conclusions:** Two doses of BioNTech-Pfizer’s BNT162b2 vaccine is highly effective against COVID-19 outcomes in elderly persons.

## Introduction

Global research on coronavirus disease (COVID-19) has shown that an increasing age is the most relevant risk factor for severe acute respiratory syndrome coronavirus 2 (SARS-CoV-2) infections.^1^ The high risk for poor outcomes of SARS-CoV-2 infection in the elderly is likely to be due to chronic comorbidities such as hypertension and cardiovascular disease,^2^ diminishing physiological functions including that of the respiratory system,^3^ and impaired immune response.^4^

The high SARS-CoV-2 infection case fatality rates in the elderly population^5^ has guided prioritization of this age group in several countries, including Germany.^6,7^ The COVID-19 disease vaccination programme in Germany was launched on Dec 27, 2020 using the BioNTech-Pfizer mRNA BNT162b2^8^ vaccine which requires two doses within at least 21 days for full protection.^9^ Other vaccines including Moderna (mRNA-1273), Oxford-AstraZeneca (ChAdOx1 nCoV-19), and Johnson & Johnson (Ad26.COV2.S) were licensed for use in Germany on Jan 6, 2021, Jan 29, 2021, and Mar 11, 2021, respectively.

In this study, we used vaccine uptake records of the large population of individuals aged ≥80 years in Bavaria (n≥700,000), Germany, to evaluate the vaccine efficacy (VE) of the BNT162b2 vaccine with respect to SARS-CoV-2 infection or COVID-19-related hospitalisation or mortality in this age group. We further assessed whether VE differed between age groups ≥80 to ≤89 years and ≥90 years.

## Methods

### Study design and population

The federal state of Bavaria is located in the South-eastern part of Germany. Of its nearly 13 million inhabitants, 831,499 (6.3%) persons were aged 80 years and above (males 38.6%, females 61.4%) by 31 Dec, 2019^10^ and were considered as reference population in the present analysis.

### The German vaccination programme

The German Ministry of Health outlined recommendations for COVID-19 vaccination in day-to-day care practices and designated vaccination centres for implementation in respective German states. All vaccines used until Apr 30, 2021 in Germany required two doses of the same vaccine, according to the Standing Committee on Vaccination (STIKO).^7^

Data on the COVID-19 vaccination was collected by authorized staff of the vaccination centers and vaccination teams and entered in the Bavarian Corona vaccination portal “BayIMCO”. During the study period cumulative vaccination data was transferred to the Robert Koch-Institut (RKI), because the transmission of records was not yet possible. The Bavarian Health and Food Safety Authority (LGL) could receive anonymous records for regional studies.

Aggregated level data on vaccination uptake in Bavaria per calendar week until Apr 11, 2021 were used for this study. Vaccination uptake data for persons aged ≥80 years included information on vaccination type (BNT162b2, ChAdOx1 nCoV-19, and mRNA-1273), number of vaccination doses received (1 or 2), sex, and age group (≥80 to <85 years, ≥85 to <90 years, ≥90 to <95 years, and ≥95 years).

### Testing for SARS-CoV-2 infection

Following the mandate given by the German Ministry of Health, notification of SARS-CoV-2 infections became obligatory since Jan 30, 2020.^11^ A positive COVID-19 case was defined as a person with a laboratory confirmation by detection of the SARS-CoV-2 virus by reverse transcription polymerase chain reaction (RT-PCR).^12,13^ Samples were tested for SARS-CoV-2 infection at the Public Health Microbiology (PHM) laboratory of the LGL,^14^ private hospitals and university laboratories, and in routine care.^13^ Further details on the Bavarian strategy of SARS-CoV-2 testing can be found elsewhere.^13^ The data were collected at the public health offices and pseudo-anonymously forwarded to the LGL.

Data on all COVID-19 cases aged ≥80 years in Bavaria between Jan 9, 2021 to Apr 11, 2021 was extracted from the LGL data bank. We used the data up until Apr 11, 2021 because as of Mar 31, 2021 vaccination was also implemented by general practitioners, who document vaccinations in a different platform and format. Thus, uniform vaccination uptake data were no longer available.

Case data included information on age, sex, whether an RT-PCR test was performed, date of infection, hospitalisation status, date and reasons of hospitalisation, mortality status, date and reasons of mortality, vaccination dose (1 or 2), as well as date and type of the last COVID-19 vaccination.

### Outcomes

We assessed VE for SARS-CoV-2 infection, COVID-19-related hospitalisation, and COVID-19-related mortality. A case with a SARS-CoV-2 infection was defined as a person with a positive RT-PCR test result. All SARS-CoV-2 infections included both symptomatic and asymptomatic cases. Hospitalisation was defined as admission to hospital with a positive RT-PCR test result. A COVID-19-related mortality was defined as a person with a positive RT-PCR test result who died because of or in temporal relation to a SARS-CoV-2 infection.

### Exclusion criteria for analysis

The analysis was limited to persons aged ≥80 years in Bavaria. We excluded observations with any of the following conditions: a) the infection was not confirmed by a positive RT-PCR test result, b) the infection was reported to have occurred before vaccination, c) the infection was related to a hospitalisation or mortality which occurred before 9th Jan, 2021, d) the vaccination was performed already in 2020, e) the person received another vaccine than BNT162b2). The exclusion criteria d) and e) applied also for the vaccine uptake dataset, with 29,071 persons already vaccinated in 2020, and 40,049 persons and 54,192 persons vaccinated with Moderna (mRNA-1273) or Oxford-AstraZeneca (ChAdOx1 nCoV-19), respectively, leaving a final number of n=708,187 for our analysis. There were no vaccinations with Johnson & Johnson (Ad26.COV2.S) recorded in the data because procurement was still outstanding during the course of the study.

### Statistical analyses

To assess VE for each outcome, we combined the information from the vaccination uptake dataset with aggregate level data and from the SARS-CoV-2 infection dataset with individual patient data. The uptake data were used to calculate the number of vaccinated and unvaccinated persons per calendar week in 2021. Persons who were vaccinated with the BNT162b2 vaccine between Jan 9, 2021 (calendar week 1) and Apr 11, 2021 (calendar week 14) contributed to the follow-up of both unvaccinated and vaccinated persons. For example, if a person was vaccinated in calendar week 5, he/she was regarded as under follow-up for 4 weeks in the unvaccinated group and for the subsequent 10 weeks in the vaccinated group, respectively (and as censored thereafter in both cases).

With the combined vaccination uptake and infection data, we calculated Kaplan-Meier (KM) plots, p-values from log-rank tests, and hazard ratios (HRs) with corresponding 95% confidence intervals (CIs) from Cox’s proportional hazard models for time to SARS-CoV-2 infection by vaccination group. Time to any SARS-CoV-2 infection, time to SARS-CoV-2 infection with later hospitalisation, and time to SARS-CoV-2 infection leading to mortality were investigated as separate outcome variables. In case the date of COVID-19 related hospitalisation or mortality was earlier than the date of confirmed SARS-CoV-2 infection, the date of SARS-CoV-2 infection was set to the date of hospitalisation or mortality, respectively. All events were inversely weighted by the number of SARS-CoV-2 infections per calendar week in the age group ≥80 years to account for temporal changes in infection risk. Having two vaccination doses was used as the main exposure variable and compared to having no vaccination, respectively. All analyses were done for the whole age group ≥80 years and the HRs were additionally stratified by age ≥80 to ≤89 years and ≥90 years. Furthermore, the KM plots were stratified by sex, and the HRs were calculated with and without adjustment for sex. VE was derived as 1 minus HR.

All analyses were carried out using R 3.6.3. The analysis code is available at https://osf.io/e9qm6/.

## Results

### Vaccine uptake in Bavaria in elderly persons

Until Apr 11, 2021, 63.8% elderly persons received one and 52.7% received the second BNT162b2 vaccination dose in Bavaria (Figure 1).

**Fig 1.**
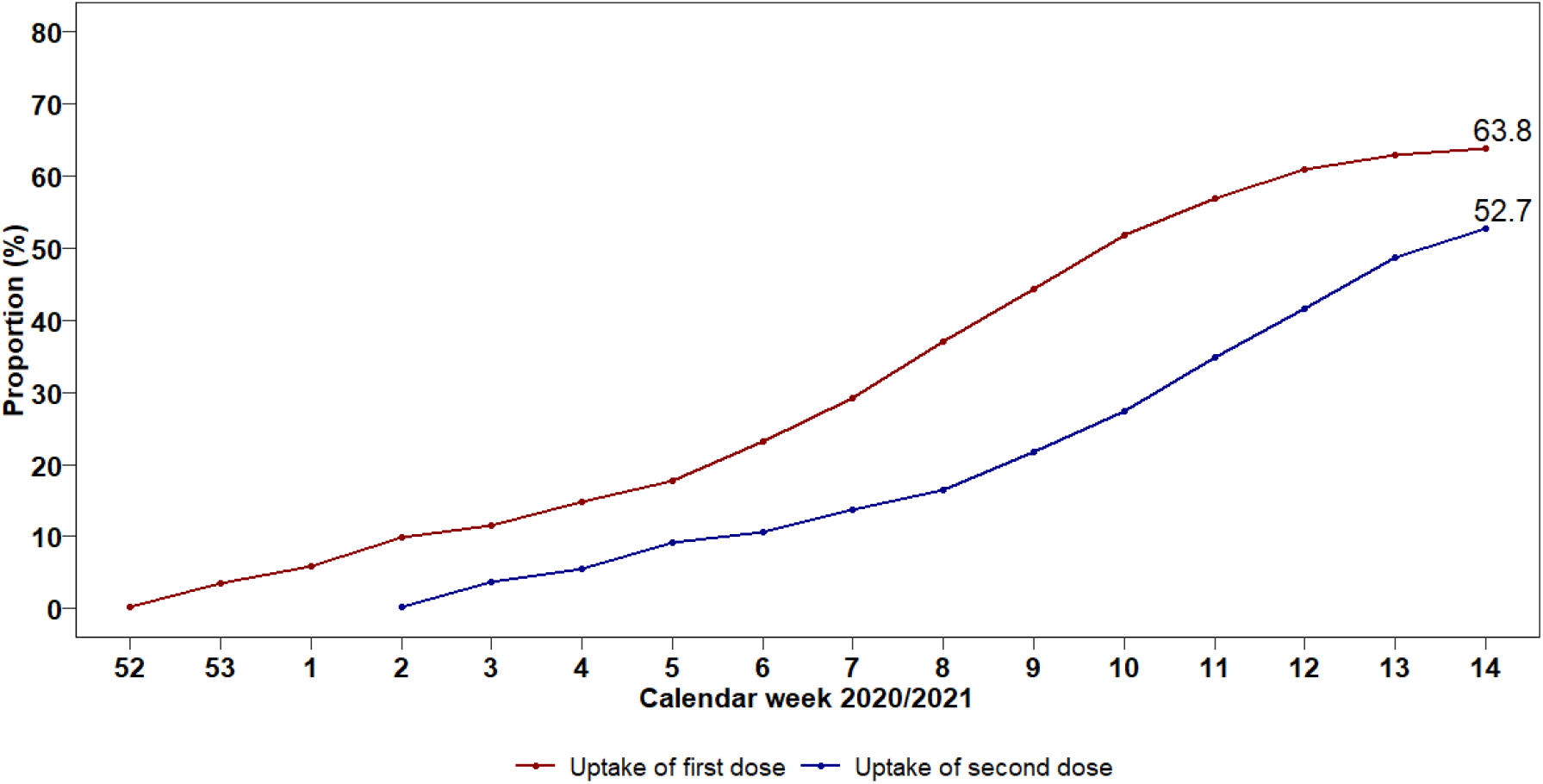
Uptake of BNT162b2 vaccine in persons ≥80 years in Bavaria.

### Confirmed COVID-19 cases in Bavaria in elderly persons

Within the follow-up period, there were 11,228 Bavarian persons aged ≥80 years with a confirmed SARS-CoV-2 infection based on a positive RT-PCR test (Fig 2). After excluding vaccinated cases with missing information on vaccine dose or type and those who were not vaccinated with the BioNTech-Pfizer BNT162b2 vaccine (n=135), a total of 1,148 vaccinated cases were considered in the vaccination group. Of these included vaccinated cases, 44.9% were partially and 55.1% were fully vaccinated with the BNT162b2 vaccine.

**Fig 2.**
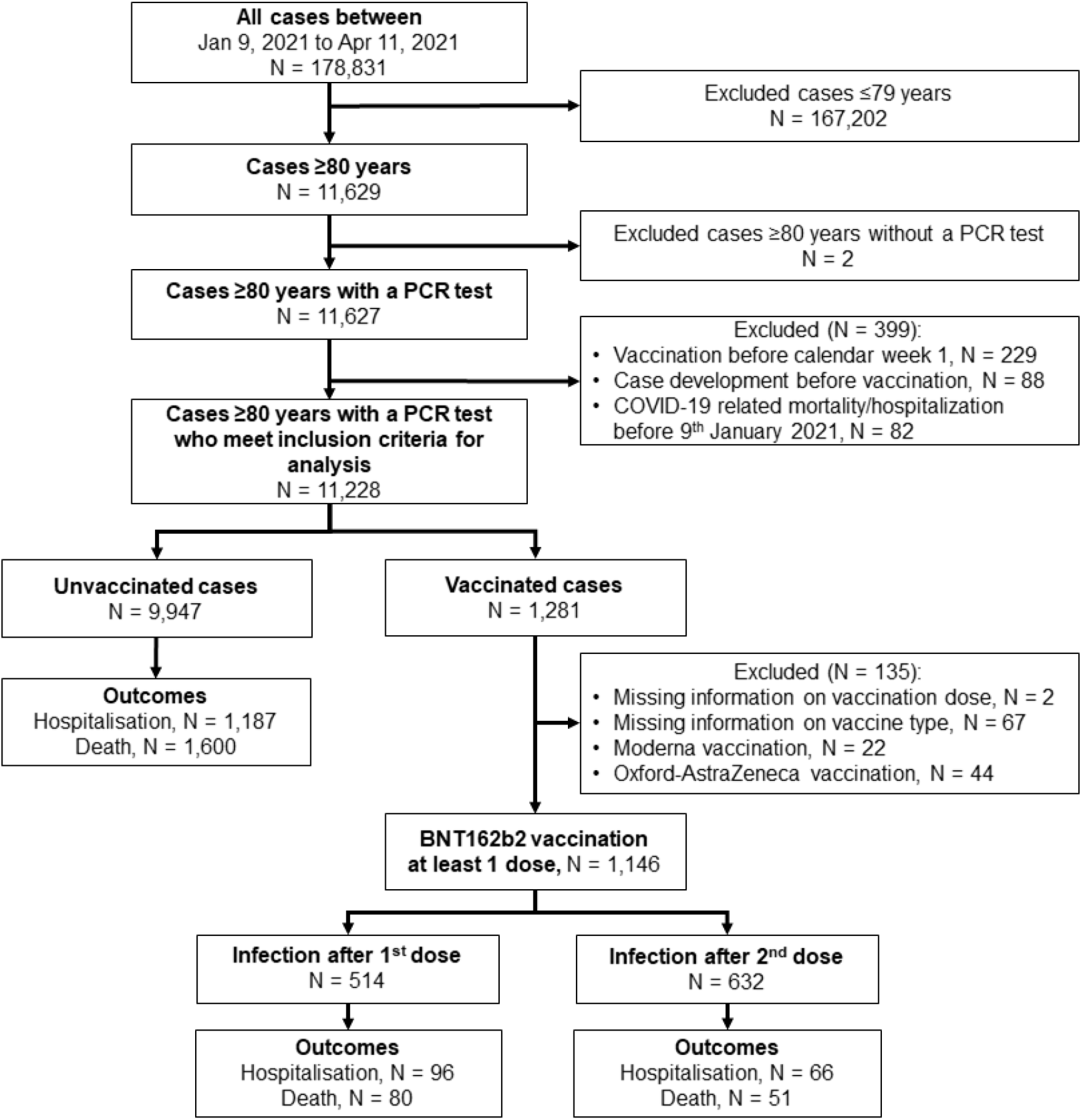
Flow chart of the study population.

The mean age of the included cases was 86.1 years (SD 4.9 years) and 35.5% were males. There were 1,349 cases who were hospitalized and 1,731 cases who died due to SARS-CoV-2 infection. The proportion of COVID-19-related hospitalisation did not differ according to vaccination status (from Chi-square test p=0.135). However, the proportion of COVID-19-related mortality was significantly lower in vaccinated persons compared to persons not vaccinated (from Chi-square test p<0.0001).

### Risk of SARS-CoV-2 infection and related outcomes

In persons aged ≥80 years, having received two compared to no BNT162b2 vaccination doses was associated with a substantially lower cumulative risk of getting a SARS-CoV-2 infection (0.41% versus 1.25% after 70 days, Figure 3A). The estimate for VE after 2 doses of the BNT162b2 vaccine to prevent SARS-CoV-2 infection was 68.3% (95% CI 65.5%, 70.9%) (Figure 4A).

**Fig 3.**
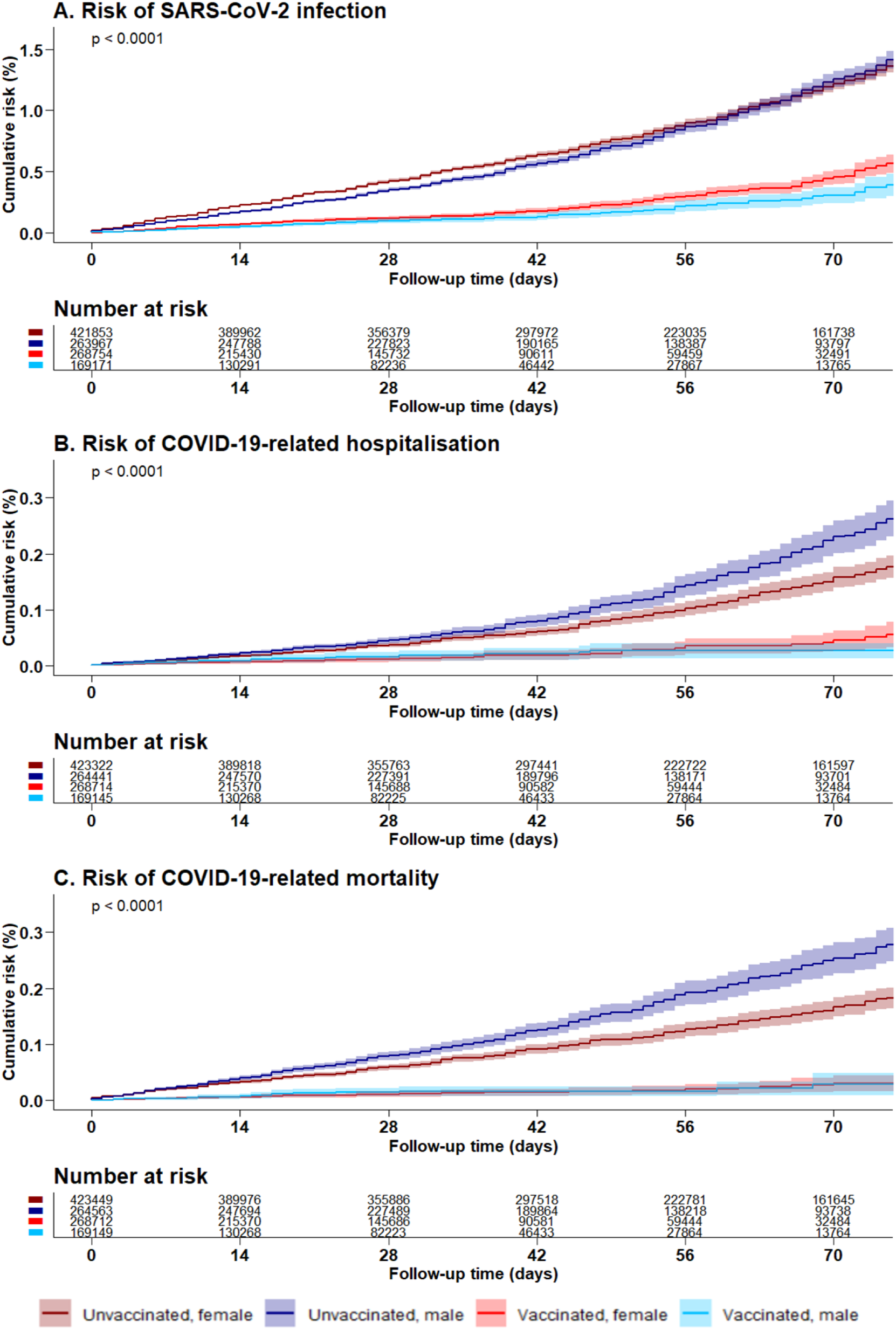
Risk of SARS-CoV-2 infection and related outcomes after two BNT162b2 vaccine doses in Bavarian persons aged 80 years and above.

**Fig 4.**
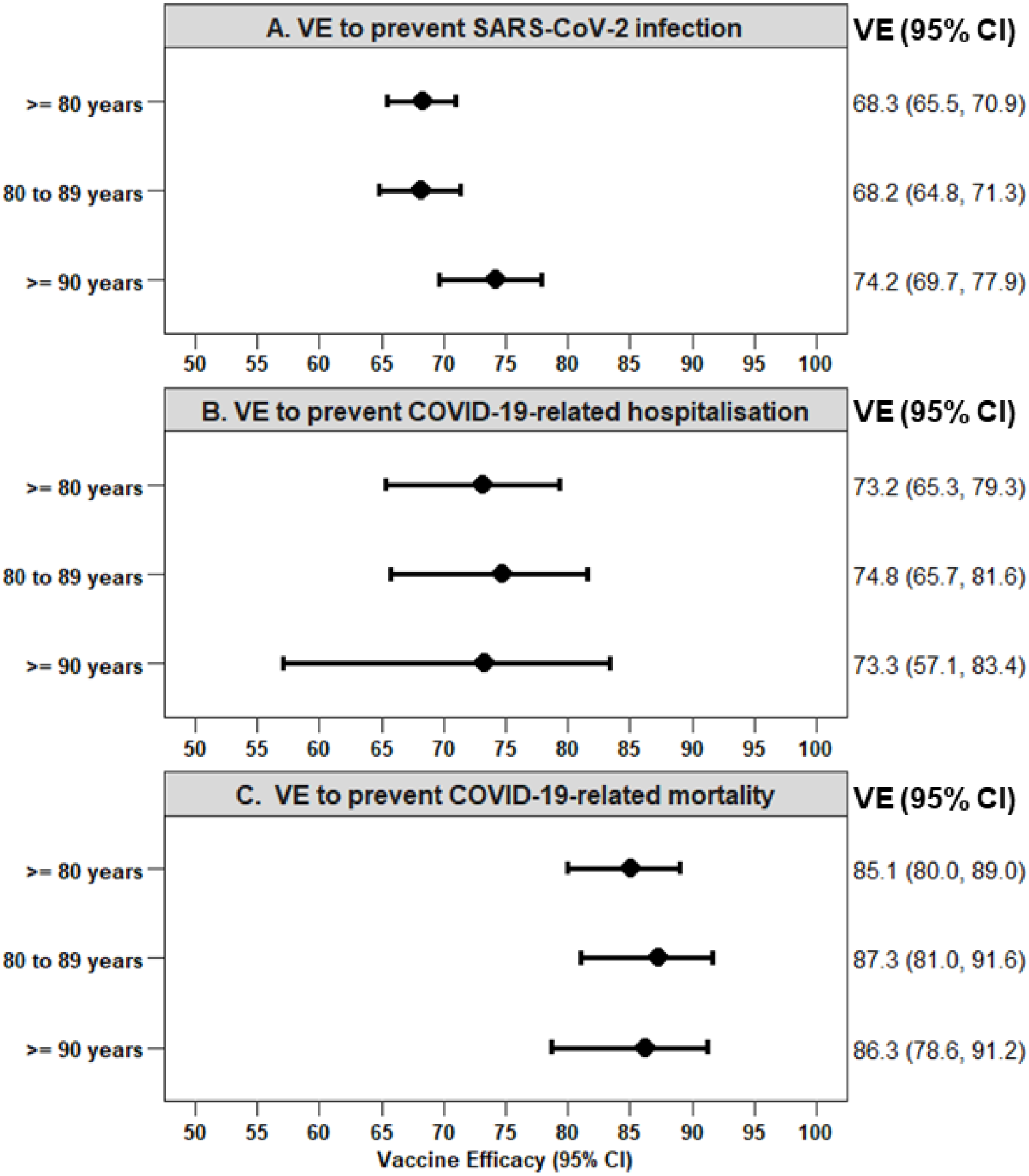
Vaccine efficacy to prevent SARS-CoV-2 infection and related outcomes after two BNT162b2 vaccine doses compared to none in Bavarian persons aged 80 years and above. Vaccine efficacy was calculated as 1 – HR, adjusted for sex. CI, confidence interval; HR, hazards ratio; VE, vaccine efficacy.

The risk of COVID-19-related hospitalisation (0.04% versus 0.19% after 70 days) and COVID-19-related mortality (0.03% versus 0.20% after 70 days) were also lower after two doses of the BNT162b2 vaccine (Figure 3B, Figure 3C). In the group of unvaccinated persons, the risk of infection was higher in females than in males and the risk for hospitalisation and mortality was higher in males than in females. After two doses of the BNT162b2 vaccine, females showed a significantly higher cumulative infection risk than males (p<0.0001 from log-rank test), while there were no significant sex differences for VE with respect to hospitalisation (p=0.91) and mortality (p=0.77), respectively.

By the end of the follow-up period, VE for COVID-19-related hospitalisation and COVID-19-related mortality was 73.2% (95% CI 65.3%, 79.3%) and 85.1% (95% CI 80.0%, 89.0%), respectively (Figure 4B, Figure 4C). Estimates of VE in octo- and novo-generians overlapped.

S1 Figure presents VE based on the HR for SARS-CoV-2 infection and related outcomes in cases with one or more doses of the BNT162b2 vaccination. The trends in risk of SARS-CoV-2 infections and related outcomes were similar after either ≥1 dose or both doses of the BNT162b2 vaccine. However, VE for all outcomes were lower in persons with at least 1 than in persons with both doses of the BNT162b2 vaccine.

## Discussion

In this population-based analysis of all Bavarian persons aged ≥80 years with suitable follow-up between Jan 9, 2021 and Apr 11, 2021, we found a high protection against SARS-CoV-2 infection and mortality after the second dose of the BNT162b2 vaccine. This association did not differ by age above ≥80 to ≤89 years and ≥90 years. In the unvaccinated group, women were more likely to be infected than men whereas men were more likely to be hospitalized or die from COVID-19. Sex differences in the risk of hospitalisation and mortality in the absence of vaccination disappeared widely after vaccination.

Population-based studies to evaluate the efficacy of COVID-19 vaccines have shown that a single dose of the BNT162b2 vaccine had an efficacy between 42% and 75% against SARS-CoV-2 infection, between 43% and 85% against COVID-19-related hospitalisation, and between 51% and 72% against COVID-19-related mortality after at least 14 days.^15^ Overall, the estimates of VE increased after the second dose.^15^ There are scarce studies addressing VE after BNT162b2 vaccination in persons 80 years and above and none distinguishing VE in octo- and novo-genarians.

In the efficacy study on the BioNTech-Pfizer trial, VE to reduce SARS-CoV-2 infection was 94.7% in persons ≥65 years and 100.0% in persons ≥75 years. However, the CI for the VE estimates included null which could be possibly due to the low proportion of elderly persons in their study. Our real life data of elderly Bavarian persons, we found an overall VE of 68.3% after two doses with small 95% confidence bands. The efficacy of 68.3% against any infection after two BNT162b2 might explain outbreaks in homes for the elderly despite high uptake of the vaccine reported in the press occasionally. Interestingly, VE did not differ between octo- and novo-genarians. The BioNTech-Pfizer efficacy study could not assess VE by outcome due to few hospitalized cases and mortality in the older age groups. Our data clearly show that VE against hospitalisation after 2 doses is the same as for any SARS-CoV-2 infection and is substantially and significantly higher for COVID-19-related mortality.

Although there was a clear difference in the risk of SARS-CoV-2 infection and COVID-19-related hospitalisation and mortality between men and women, no such difference was observed after vaccination for hospitalisation or mortality, suggesting that BNT162b2 is equally effective for severe outcomes in elderly men and women.

An important study on elderly persons has addressed the onset of effect after one and two doses of the BNT162b2 or the Oxford-AstraZeneca vaccine.^16^ The study showed that the VE increases up to 42 days after the first dose and up to after 14 days after the second dose. While the risk for infection decreased only gradually within the first 42 days after the first dose, VE after the second dose already reached 80% after 7 days. During vaccination programmes requiring two doses, it is evident that some persons will only be vaccinated once up to the second dose. In order to estimate the effect of the vaccination programme, it may also be interesting to know the effect of one or more doses. In a population where most second doses were given within 3 weeks after the first dose, VE was found only slightly lower after 2 doses with overlapping 95% CIs.

A strength of our study is the large sample of elderly persons living in Bavaria. Data was retrieved from government data bases and have been checked for implausibilities. Given the structure of the data base for cases which only included date of the last vaccination and number of vaccine doses, we were unfortunately unable to evaluate the VE after the first BNT162b2 vaccine dose. Further, information of risk factors which could contribute to severe COVID-19 disease outcomes in persons aged 80 years and above have not been ascertained for the cases.

In conclusion, we found that two doses of BioNTech-Pfizer’s BNT162b2 vaccine provided a high protection against SARS-CoV-2 infection and severe outcomes in elderly persons. There was no difference between octo- and novo-generians and efficacy appeared to be equal in both sexes.

## Data Availability

Data cannot be shared publicly because participants did not explicitly consent to the sharing of their data as per European Unions General Data Protection Regulation and the corresponding German privacy laws. Data are available through the Research Ethics Board of the Ludwig-Maximilians-Universitaet Muenchen, Munich/Germany for researchers who meet the criteria for access to confidential data.

## Abbreviations

CI: confidence interval
COVID-19: coronavirus disease
HR: hazard ratios
KM: Kaplan-Meier
LGL: Bavarian Health and Food Safety Authority
PHM: Public Health Microbiology
RKI: Robert Koch-Institute
RT-PCR: reverse transcription polymerase chain reaction
SARS-CoV-2: severe acute respiratory syndrome coronavirus 2
STIKO: Standing Committee on Vaccination
VE: vaccine efficacy

## Acknowledgements

We are grateful to the public health offices in Bavaria, Germany who supplied data on cases with SARS-CoV-2 infection and to teams of the Bavarian Corona vaccination portal “BayIMCO” for providing the COVID-19 vaccination data.

## Author Contributions

**Conceptualization:** Klaus Überla, Rüdiger von Kries

**Data Curation:** Delphina Gomes, Andreas Beyerlein, Katharina Katz, Gabriele Hölscher, Uta Nennstiel, Bernhard Liebl

**Formal Analysis:** Andreas Beyerlein, Delphina Gomes

**Funding Acquisition:** Klaus Überla, Rüdiger von Kries

**Investigation:** Rüdiger von Kries

**Methodology:** Andreas Beyerlein, Rüdiger von Kries

**Project Administration:** Rüdiger von Kries

**Software:** Andreas Beyerlein, Delphina Gomes

**Supervision:** Rüdiger von Kries

**Validation:** Andreas Beyerlein, Delphina Gomes, Rüdiger von Kries

**Visualization:** Andreas Beyerlein, Delphina Gomes, Rüdiger von Kries

**Writing – Original Draft Preparation:** Delphina Gomes, Andreas Beyerlein, Rüdiger von Kries

**Writing – Review & Editing:** Delphina Gomes, Andreas Beyerlein, Katharina Katz, Gabriele Hölscher, Uta Nennstiel, Bernhard Liebl, Klaus Überla, Rüdiger von Kries

## Supporting information

**S1 Figure.**
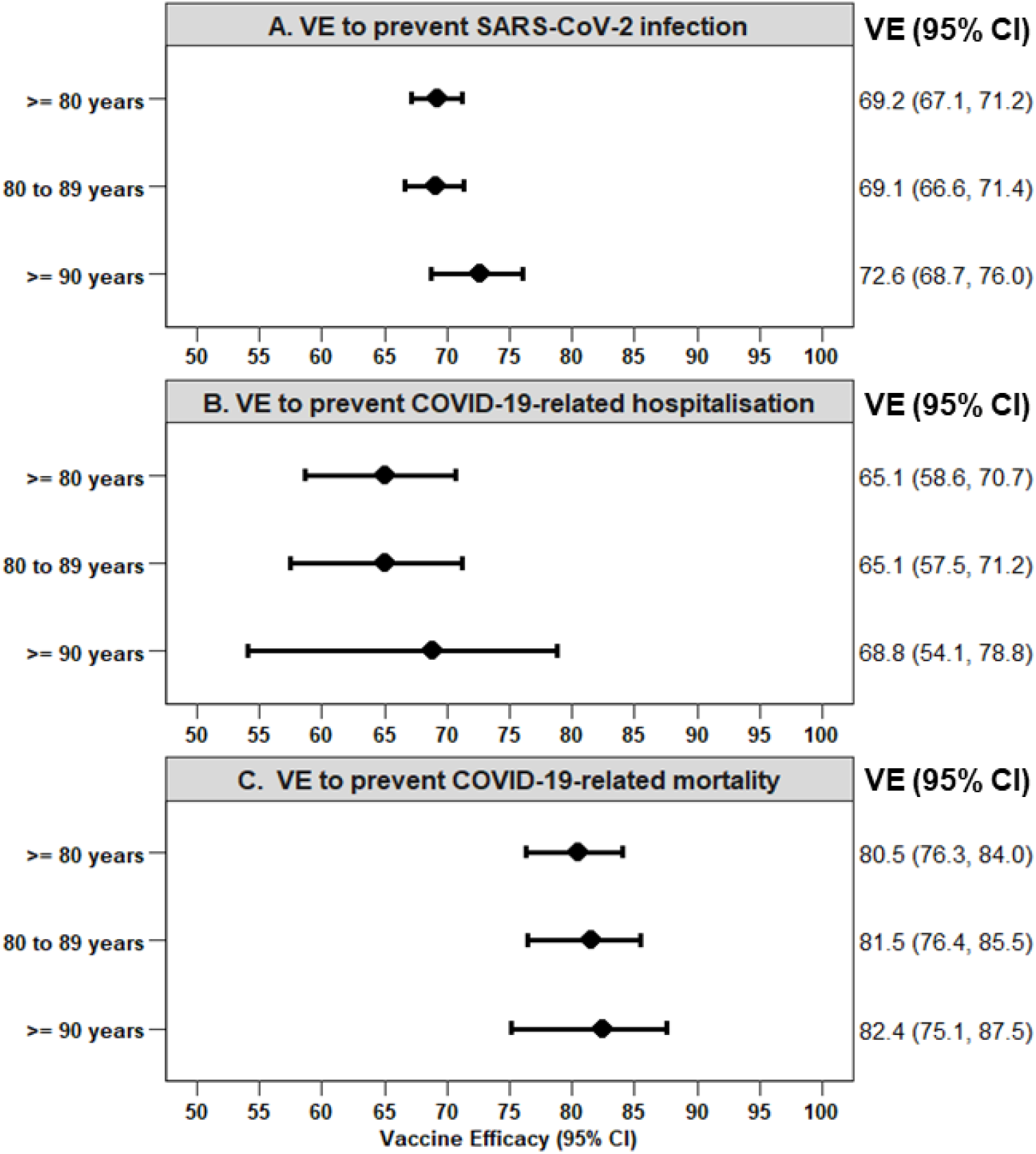
Vaccine efficacy to prevent SARS-CoV-2 infection and related outcomes after at least one BNT162b2 vaccine dose compared to none in Bavarian persons aged 80 years and above. Vaccine efficacy was calculated as 1 – HR and are adjusted for sex. CI, confidence interval; HR, hazards ratio; VE, vaccine efficacy.

