## Supplementary material for "Is the BioNTech-Pfizer COVID-19 vaccination effective in elderly populations? Results from population data from Bavaria, Germany": S1 Figure.

**S1 Figure. Vaccine efficacy to prevent COVID-19 infection and related outcomes after at least one BNT162b2 vaccine dose compared to none in Bavarian persons aged 80 years and above.**

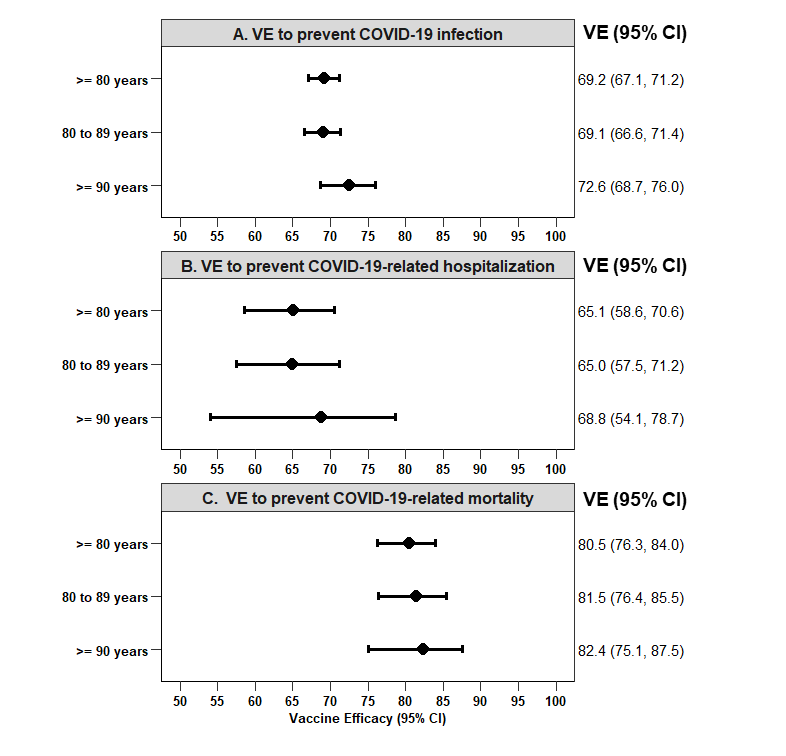
Vaccine efficacy was calculated as 1 – HR and are adjusted for sex. CI, confidence interval; HR, hazards ratio; VE, vaccine efficacy.
